# Treatment of obesity in US children and adolescents before and after the AAP guidelines

**DOI:** 10.1101/2024.10.25.24316147

**Authors:** Patricia J Rodriguez, Duy Do, Samuel Gratzl, Brianna M Goodwin Cartwright, Nicholas L Stucky, Davene Wright R.

## Abstract

**Importance:** The American Academy of Pediatrics (AAP) published new comprehensive guidelines for childhood obesity treatment, including pharmacotherapy. However, changes in treatment patterns following these guidelines remain unknown.

**Objective:** To assess changes in childhood and adolescent obesity treatment following the new AAP guidelines.

**Design:** Using Truveta electronic health record (EHR) data, this cohort study analyzed outpatient visits between January 2021 and June 2024 for children (age 8-11) and adolescents (age 12-17) with obesity and no evidence of type 2 diabetes (T2D). For patients with multiple visits, one visit was randomly selected. Patients without a recent history of specific obesity treatments were followed for evidence of obesity treatment at or following their visit, including nutrition referral within 14 days or nutrition counseling within 90 days, and, separately, pharmacotherapy prescriptions for weight management (on- or off-label) within 14 days. Interrupted time series models were used to compare differences in nutrition counseling or referral and pharmacotherapy before versus after the AAP guidelines were released.

**Setting:** Clinical and prescribing data from EHRs from a collective of US health systems.

**Participants:** 329,357 patients aged 8 to 17 with an outpatient office visit, a BMI percentile indicating obesity, and no T2D diagnosis.

**Exposure:** Release of the AAP guidelines in January 2023.

**Main Outcomes and Measures:** Evidence of obesity treatment at or following the eligible visit with (1) nutrition referral or counseling, and (2) pharmacotherapy.

**Results:** The study population of 329,581 patients included 120,734 (36.6%) children and 208,847 (63.4%) adolescents. The mean (SD) BMI percentile was 97.4 (1.6), with 119,864 (36.4%) having severe obesity (class 2 or 3). Overall, a minority of patients without a recent history of obesity treatment had evidence of nutrition referral or counseling (4.0%) or pharmacotherapy (0.4%) during or shortly after their visit. Following the AAP guidelines, indicators of both immediate (odds ratio [95% CI]: 1.38 [1.08-1.75]) and gradual monthly changes (1.06 [1.04-1.08]) were observed for pharmacotherapy use. An immediate change in nutrition counseling and referral was observed following guidelines (1.34 [1.24-1.45]), but no additional gradual monthly changes were observed (1.0 [0.99-1.01]).

**Conclusions and Relevance:** While nutrition counseling or referral and pharmacotherapy use increased, overall rates of obesity treatment remained low.

## Background

Obesity occurs in approximately 20% of youth in the US,^1^ and is associated with increased short- and long-term health risks, including cardiovascular disease, type 2 diabetes (T2D), hypertension, hyperlipidemia, and mental health conditions.^2–5^ In January 2023, the American Academy of Pediatrics (AAP) issued its first recommendations for the evaluation and treatment of obesity among children and adolescents, which emphasized the importance of early, evidence-based treatment.^6^

The AAP guidelines identify intensive health behavior and lifestyle treatment (IHBLT) as the first-line intervention. This approach involves frequent, family-based counseling with highly trained providers over several months, ideally delivered in a face-to-face setting for a total of at least 26 hours.^6^ Lifestyle intervention alone has shown modest weight reductions for youth, and access to intensive programs is often limited, particularly outside large children’s hospitals in urban areas.^7,8^ For adolescents aged 12 and older, the AAP guidelines also recommend offering pharmacotherapy as an adjunct to IHBLT.^6^ The guidelines do not recommend pharmacotherapy for children aged 8-11given insufficient evidence, though it may be considered on a case-by-case basis.

The fact that AAP guidelines supported early pharmacotherapy was met with some criticism.^9^ Concomitant with the rising popularity of new, highly effective glucagon-like peptide-1 receptor agonists (GLP-1 RA) in the US,^10^ concerns have been raised about inappropriate overuse of pharmacotherapy in children, weight stigmatization, and the risk of eating disorders.^11^ In addition, GLP-1 RA use in adolescents is likely not cost-effective at current prices.^12^ While previous studies have identified minimal use of pharmacotherapy in children and adolescents,^13,14^ little is known about use following the AAP guideline release.

Given these concerns, this real-world retrospective cohort study aims to evaluate whether the use of treatments for obesity for children and adolescents, including nutrition counseling and pharmacotherapy, changed after the release of the AAP guideline.

## Methods

### Study Design

This study utilized electronic health records (EHR) from a group of US health systems to identify visits for children and adolescents aged 8-17 that were potentially eligible for obesity treatment. Eligible visits were defined as outpatient office visits where the patient had a body mass index (BMI) meeting obesity criteria (defined below) and no evidence of recent specific obesity treatment. Patients were followed to assess obesity treatment during or shortly after the visit, including referral to a nutritionist or dietician, evidence of nutritional/dietary counseling, and prescriptions of pharmacotherapies used for weight management. Interrupted time series (ITS) models were used to compare the likelihood of receiving obesity treatments before versus after the AAP guidelines were released.

### Data

This study used a subset of Truveta Data, which provides access to continuously updated and linked EHR data from a collective of US health care systems. Data include structured information on demographics (age, sex, health system-reported race and ethnicity), encounters, diagnoses, vital signs (e.g., weight, height, BMI), medication requests (prescriptions), laboratory tests and results (e.g., HbA1c), and procedures.

Data were normalized into a common data model through syntactic and semantic normalization. Truveta Data were then de-identified by expert determination under the HIPAA Privacy Rule. This study used only de-identified patient records and therefore did not require Institutional Review Board approval. Data for this study were accessed on October 17, 2024 using Truveta Studio.

### Study population and exposure

Visits for children (ages 8-11) and adolescents (ages 12-17) potentially eligible for obesity treatment were identified. Eligible visits included outpatient and office visits between January 2021 and June 2024, where a patient’s BMI met obesity criteria and the patient had no previous diagnosis of T2D. For patients with multiple qualifying visits, a single visit was randomly selected as the index visit. Obesity was defined as BMI greater than the 95^th^ percentile for age (in months) and sex, as defined by the CDC.^15^ Obesity class was similarly defined using CDC criteria. BMIs of 120% to less than 140% of the 95^th^ percentile or 35 to less than 40 kg/m^2^ were classified as class II obesity, while those 140% or more of the 95^th^ percentile or 40 kg/m^2^ or greater were classified as class III obesity.^16^

The AAP guidelines were released on January 7, 2023. For patient privacy reasons, exact visit dates in these data were obscured to the first day of the month. Any visits occurring before January 1, 2023 were considered pre-guideline, while any visits occurring after January 1, 2023 were considered post-guideline. For all post-guideline visits, the number of months elapsed since guideline release were also defined.

Patient comorbidities, treatment history, and prior medication prescribing and dispensing were characterized at the time of the index visit. Comorbidities, including ADHD, asthma, hyperlipidemia, hypertension, and obstructive sleep apnea (OSA) were identified, considering all available history up to and including the index visit. Recent treatment (nutrition counseling or referral and pharmacotherapy) was assessed in the year prior to the index visit. Previous evidence of pharmacotherapy included dispensing information, which is not limited to medication prescribed within Truveta-constituent health systems.

### Outcomes of obesity treatment

This study separately considered two obesity treatments: (1) nutrition counseling or referral, and (2) pharmacotherapies. Patients with a recent history of the specific treatment (in the previous year) were excluded from treatment-specific analyses. A one-year period was selected as the recent treatment period given the expectation of approximately annual child health visits, a recommendation for at least 26 weeks of IHBLT, and the need to wait for intervention effects to be observed given rapid changes in height and weight with puberty.

#### Nutrition counseling

Nutrition counseling and referrals for nutrition counseling served as a proxy for IHBLT, the primary, first-line treatment recommended by the AAP guidelines.^6^ Given delays between visits and referrals or counseling, we considered outcomes of nutrition referrals within 14 days of the eligible visits and counseling within 90 days. Nutrition counseling was identified using SNOMED-CT, ICD-10, and HCPCS codes for nutrition/diet counseling, education, nutrition care education, nutrition assessment, nutrition and dietary counseling and surveillance. SNOMED-CT codes were used to identify referrals to dieticians and nutrition professionals. Codes are provided in supplemental tables 1 and 2.

#### Pharmacotherapy

Pharmacotherapy outcomes were identified as medication requests (e.g., prescriptions written) for pharmacotherapies listed in the AAP guidelines within 14 days of the visit, including on- or off-label use of metformin, GLP-1 RA medications, orlistat, phentermine alone, topiramate alone, and combination phentermine-topiramate.^6,17,18^ Lisdexamfetamine, which was included in the AAP guidelines due to an indication for binge-eating disorder, was not included because it is predominantly used for ADHD, which may have been affected by supply disruptions of other ADHD medications during the study period.^19^ Furthermore, this analysis included all GLP-1-based drugs, without restriction to approved use or population. Only liraglutide (as Saxenda) and semaglutide (as Wegovy) are approved for weight management in adolescents. Dulaglutide and exenatide are approved for adolescents with T2D, but may be used off-label in patients without T2D.^14^ Other GLP-1-based medications, including the dual agonist tirzepatide, are approved for use in adults only, but may be used off-label in younger populations.^14^ For brevity, we use the term GLP-1 to refer to both GLP-1 RA and dual agonists.

A sensitivity analysis was performed focusing exclusively on adolescents, given the AAP pharmacotherapy recommendation was specific to patients aged 12 and older (supplement).

### Statistical analysis

Using logistic regression, separate interrupted time series models assessed the impact of the AAP guideline release on the receipt of treatment with (1) nutrition referral or counseling and (2) pharmacotherapy. All models controlled for patient demographics (age, age group [child vs adolescent], sex, race), BMI percentile at the visit, and clinician specialty. A state fixed-effect was used to account for clustering within health systems.

Two primary effects were considered: (1) a level change in the intercept after the AAP guideline release, which represents an immediate change in the likelihood of receiving treatment, and (2) a slope change after the AAP guideline release, which represents a more gradual monthly change in the use of treatment.

## Results

### Patient characteristics

During the study period, there were 3,633,285 outpatient office visits with BMI information from 1,336,610 patients without T2D. Of these, 839,043 visits (23.1%) from 329,581 (24.7%) patients met the obesity criteria. A single visit was selected randomly per patient, yielding a final study population of 329,581visits from the same number of patients.

The study population included 120,734 (36.6%) children (ages 8-11) and 208,847 (63.4%) adolescents (ages 12-17). The mean (SD) age overall was 13.2 (3.0) and 150,390 (45.6%) patients were female. Based on health system documented race in the EHR, 3,488 (1.1%) patients were American Indian or Alaska Native, 12,224 (3.7%) were Asian, 49,419 (15.0%) were Black or African American, 177,328 (53.8%) were White, 39,725 (12.1%) were other races, and 47,397 (14.4%) had unknown race. The mean (SD) BMI percentile was 97.4 (1.6), with 209,717 (63.6%) having class 1 obesity, 77,834 (23.6%) having class II obesity, and 42,030 (12.8%) having class III obesity.

Clinician background data was not available for all visits. Of visits where clinician background information was available (155,215 [47.1%]), most visits (135,272 [87.2%]) occurred with clinicians traditionally involved in primary health care delivery, including pediatricians (61,452 [39.6%]), family or internal medicine practitioners (29,755 [19.2%]), physician assistants or nurse practitioners (30,294 (19.5%]) and residents (19,964 [12.9%]).

The pre-guideline period included 184,126 patients and the post-guideline period included 145,455 patients. Demographic and health characteristics at patient visits were largely similar between these two periods, although patients in the post-guideline period were slightly younger on average (Table 1).

**Table 1:** Patient characteristics. Continuous variables displayed as mean (SD). Categorical variables displayed as number (column percent). Other race includes multiple races, Native Hawaiian, Pacific Islander, and other single races. Other state includes all states representing less than 1% of the study population.

|  | PRE-GUIDELINE<br>N = 184,126 | POST-GUIDELINE<br>N = 145,455 | OVERALL<br>N = 329,581 |
| --- | --- | --- | --- |
| Age Group |  |  |  |
| Adolescent (ages 12-17) | 121,870 (66.2%) | 86,977 (59.8%) | 208,847 (63.4%) |
| Child (ages 8-11) | 62,256 (33.8%) | 58,478 (40.2%) | 120,734 (36.6%) |
| Age | 13.4 (2.9) | 13.0 (3.0) | 13.2 (3.0) |
| Female | 82,881 (45.0%) | 67,509 (46.4%) | 150,390 (45.6%) |
| Race |  |  |  |
| American Indian or Alaska Native | 1,969 (1.1%) | 1,519 (1.0%) | 3,488 (1.1%) |
| Asian | 6,724 (3.7%) | 5,500 (3.8%) | 12,224 (3.7%) |
| Black* | 28,933 (15.7%) | 20,486 (14.1%) | 49,419 (15.0%) |
| White | 98,367 (53.4%) | 78,961 (54.3%) | 177,328 (53.8%) |
| Other | 22,578 (12.3%) | 17,147 (11.8%) | 39,725 (12.1%) |
| Unknown | 25,555 (13.9%) | 21,842 (15.0%) | 47,397 (14.4%) |
| Ethnicity |  |  |  |
| Hispanic or Latino | 43,092 (23.4%) | 33,512 (23.0%) | 76,604 (23.2%) |
| Not Hispanic or Latino | 118,474 (64.3%) | 92,546 (63.6%) | 211,020 (64.0%) |
| Unknown | 22,560 (12.3%) | 19,397 (13.3%) | 41,957 (12.7%) |
| State |  |  |  |
| Illinois | 27,708 (15.0%) | 19,728 (13.6%) | 47,436 (14.4%) |
| New York | 21,466 (11.7%) | 17,239 (11.9%) | 38,705 (11.7%) |
| Washington | 19,533 (10.6%) | 14,917 (10.3%) | 34,450 (10.5%) |
| Wisconsin | 17,996 (9.8%) | 13,044 (9.0%) | 31,040 (9.4%) |
| Michigan | 14,934 (8.1%) | 12,255 (8.4%) | 27,189 (8.2%) |
| California | 14,578 (7.9%) | 9,689 (6.7%) | 24,267 (7.4%) |
| Ohio | 13,911 (7.6%) | 8,825 (6.1%) | 22,736 (6.9%) |
| Texas | 9,374 (5.1%) | 10,092 (6.9%) | 19,466 (5.9%) |
| Virginia | 7,386 (4.0%) | 6,072 (4.2%) | 13,458 (4.1%) |
| Oregon | 6,162 (3.3%) | 4,829 (3.3%) | 10,991 (3.3%) |
| Arizona | 3,689 (2.0%) | 4,027 (2.8%) | 7,716 (2.3%) |
| New Jersey | 3,264 (1.8%) | 4,405 (3.0%) | 7,669 (2.3%) |
| Other | 6,906 (3.8%) | 10,132 (7.0%) | 17,038 (5.2%) |
| Unknown | 17,219 (9.4%) | 10,201 (7.0%) | 27,420 (8.3%) |
| BMI Percentile | 97.4 (1.6) | 97.5 (1.6) | 97.4 (1.6) |
| BMI Value | 30.6 (5.9) | 30.1 (5.9) | 30.4 (5.9) |
| BMI Class |  |  |  |
| Class 1 obesity | 117,000 (63.5%) | 92,717 (63.7%) | 209,717 (63.6%) |
| Class 2 obesity | 43,493 (23.6%) | 34,341 (23.6%) | 77,834 (23.6%) |
| Class 3 obesity | 23,633 (12.8%) | 18,397 (12.6%) | 42,030 (12.8%) |
| Severe obesity | 67,126 (36.5%) | 52,738 (36.3%) | 119,864 (36.4%) |
| Clinician Background |  |  |  |
| Pediatrics | 34,799 (18.9%) | 26,845 (18.5%) | 61,644 (18.7%) |
| Family or Internal Medicine | 17,508 (9.5%) | 12,309 (8.5%) | 29,817 (9.0%) |
| PA/NP | 16,339 (8.9%) | 13,957 (9.6%) | 30,296 (9.2%) |
| Residency | 6,570 (3.6%) | 6,919 (4.8%) | 13,489 (4.1%) |
| Other or unknown | 108,910 (59.1%) | 85,425 (58.7%) | 194,335 (59.0%) |
| History of nutrition counseling or referral |  |  |  |
| Previous 30 days | 883 (0.5%) | 775 (0.5%) | 1,658 (0.5%) |
| Previous 90 days | 2,928 (1.6%) | 2,670 (1.8%) | 5,598 (1.7%) |
| Previous year | 10,579 (5.7%) | 9,898 (6.8%) | 20,477 (6.2%) |
| History of anti-obesity medications |  |  |  |
| Previous 30 days | 715 (0.4%) | 610 (0.4%) | 1,325 (0.4%) |
| Previous 90 days | 1,317 (0.7%) | 1,198 (0.8%) | 2,515 (0.8%) |
| Previous year | 2,336 (1.3%) | 2,144 (1.5%) | 4,480 (1.4%) |
| Comorbidities |  |  |  |
| ADHD | 18,339 (10.0%) | 15,510 (10.7%) | 33,849 (10.3%) |
| Asthma | 34,041 (18.5%) | 26,771 (18.4%) | 60,812 (18.5%) |
| Hypertension | 3,451 (1.9%) | 2,509 (1.7%) | 5,960 (1.8%) |
| Hyperlipidemia | 8,492 (4.6%) | 6,373 (4.4%) | 14,865 (4.5%) |
| Obstructive sleep apnea | 7,350 (4.0%) | 7,245 (5.0%) | 14,595 (4.4%) |
| Utilization (count in previous year) |  |  |  |
| A1c test | 0.1 (0.4) | 0.1 (0.3) | 0.1 (0.3) |
| OP office visits | 1.7 (3.7) | 1.8 (3.6) | 1.7 (3.7) |
| BMI measures | 2.6 (1.0) | 2.6 (1.0) | 2.6 (1.0) |

### Nutrition referral or counseling

Of the 329,581 patients in this study, 20,477 (6.2%) had evidence of recent nutrition referral or counseling in the year before their visit and were excluded from nutrition treatment analyses. Of the remaining 309,104 patients, 12,457 (4.0%) had evidence of nutrition referral within 14 days of the visit or nutrition counseling within 90 days after the visit. Likelihood of nutrition counseling or referral was higher in patients with Asian and Black or African American race, compared to White race, and higher for visits with a pediatrician, compared to other clinician backgrounds (Figure 1).

**Figure 1:**
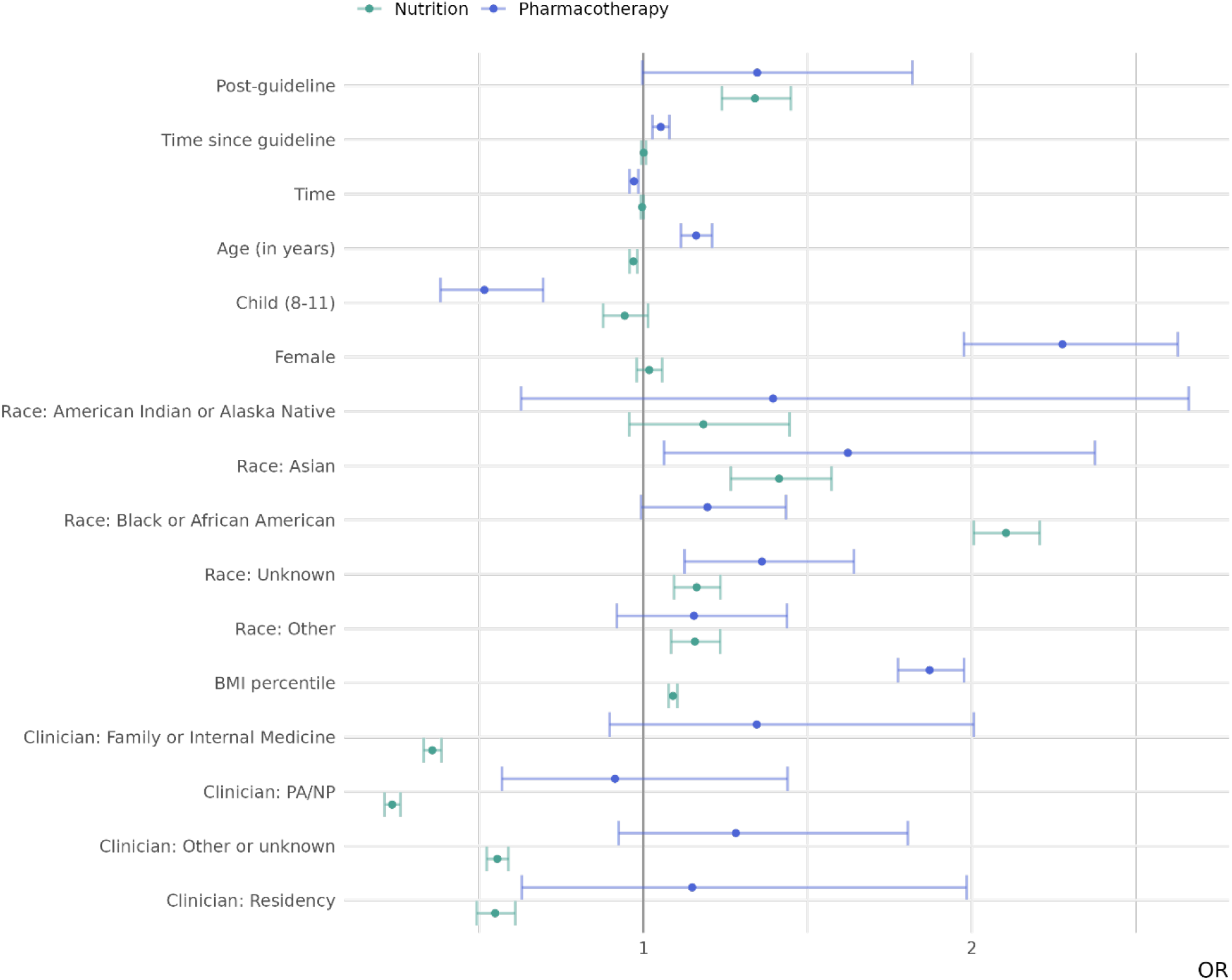
Selected adjusted odds ratios from separate logistic regressions considering the probability of treatment before and after guidelines with each intervention. Points represent adjusted odds ratios and lines represent 95% confidence intervals. Patients with a recent history (in the prior year) of the specific treatment were excluded. ORs for race are relative to white and ORs for clinicians are relative to pediatricians. Time represents time (in months) since study start and time since guideline represents time (in months) since the AAP guideline release for observations in the post-guideline period.

A significant increase was observed in the likelihood of nutrition counseling or referral (intercept) immediately after the AAP guideline release (odds ratio [95% CI]: 1.34 [1.24-1.45]). However, the monthly change in treatment rate (change in slope) during the post-guideline period was not significantly different from the pre-guideline period (OR: 1.0 [0.99-1.01]).

### Pharmacotherapy

Of the 329,581 patients in this study, 4,480 (1.4%) had evidence of pharmacotherapy prescribing or dispensing in the year before the visit and were excluded from pharmacotherapy treatment analyses. Of the remaining 325,101 patients, 1,333 (0.4%) had evidence of pharmacotherapy prescribing within 14 days after the index visit. Prescribing rates were higher among female patients, those with higher BMI, and adolescents (Figure 1).

Significant increases were observed in both the prescribing level (intercept) (OR: 1.38 [1.08-1.75]) and monthly prescribing trend (slope) (OR: 1.06 [1.04-1.08]) following the AAP guideline release (Figure 3). Results of the sensitivity analysis including adolescents only were similar to the primary analysis (supplementary Figure 1).

**Figure 2:**
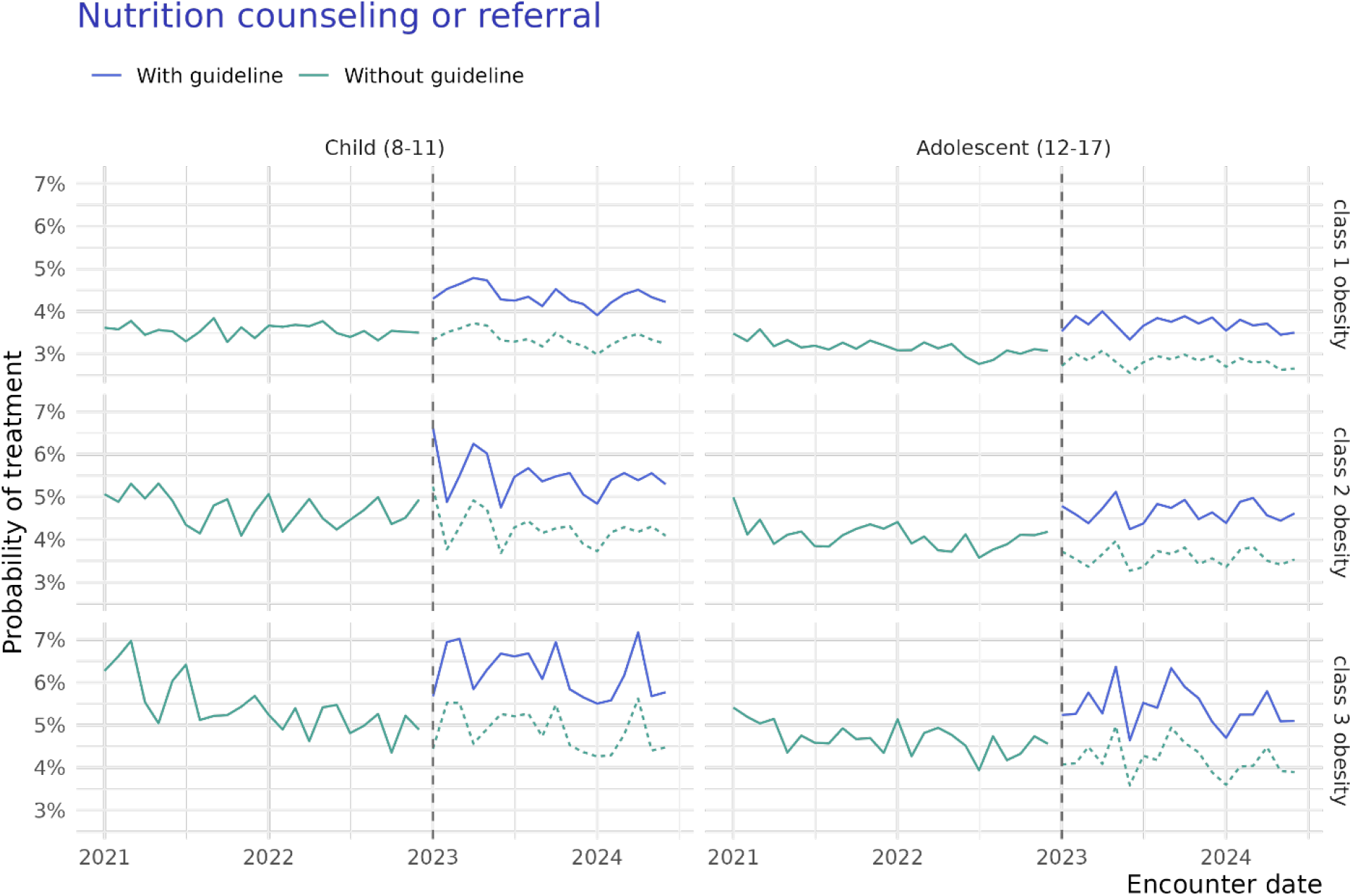
Interrupted time series of treatment with nutrition counseling or referral over time, by age and BMI class. Green lines represent the predicted probability of treatment absent guidelines, and blue lines represent predicted probabilities of treatment with the guideline, for the observed population. The dotted green line represents the counterfactual.

**Figure 3:**
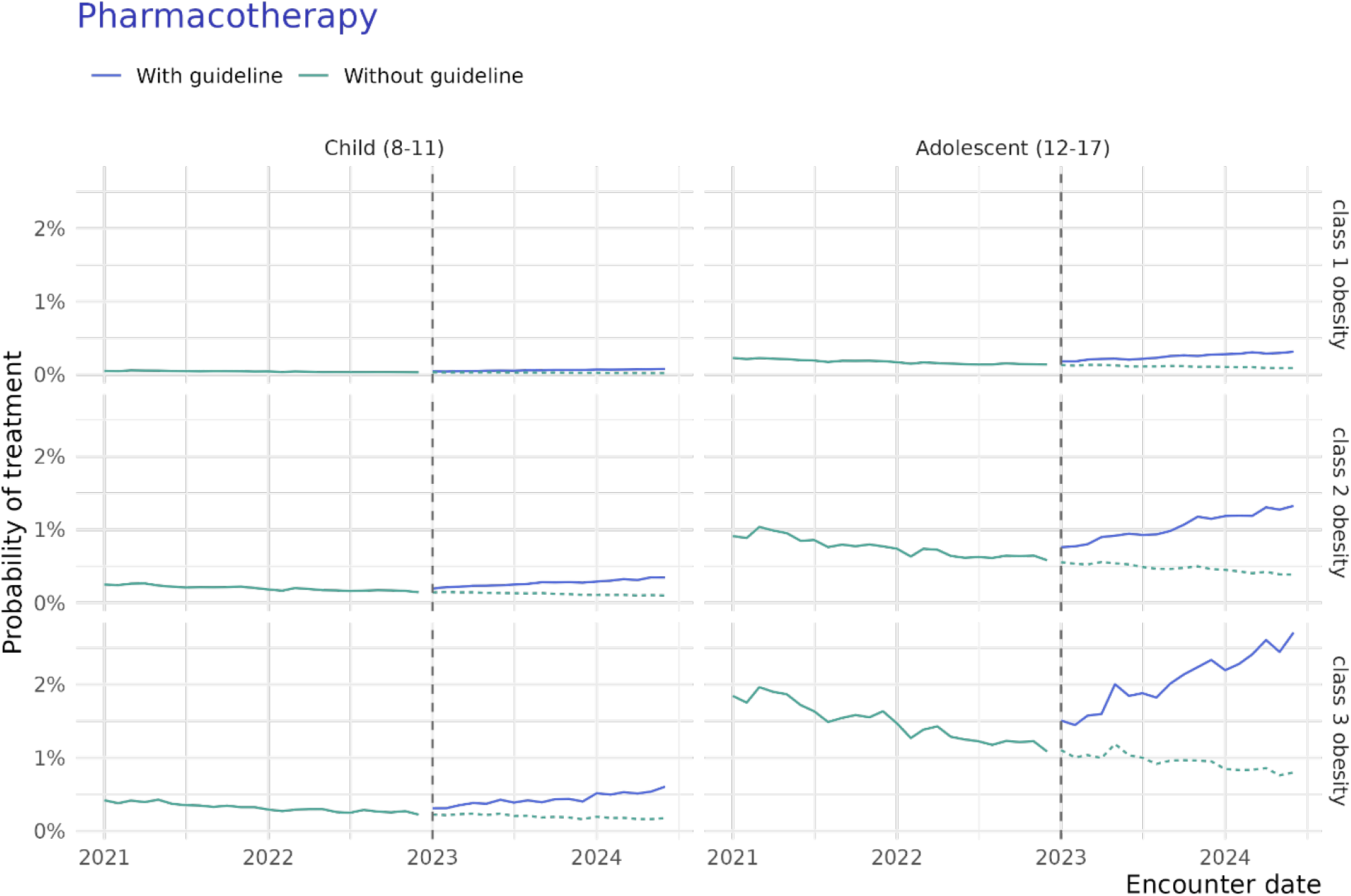
Interrupted time series of treatment with anti-obesity medication over time, by age and BMI class. Green lines represent the predicted probability of treatment absent guidelines, and blue lines represent predicted probabilities of treatment with the guideline, for the observed population. The dotted green line represents the counterfactual.

Metformin was the most commonly prescribed medication (Figure 4). During the pre-guideline period, 81.9% of patients newly prescribed pharmacotherapy were prescribed metformin, while fewer than 2% were prescribed semaglutide (prescriptions are not mutually exclusive). In contrast, during the post-guideline period, 68.5% of patients newly prescribed pharmacotherapy were prescribed metformin, while 20.0% were prescribed semaglutide.

**Figure 4:**
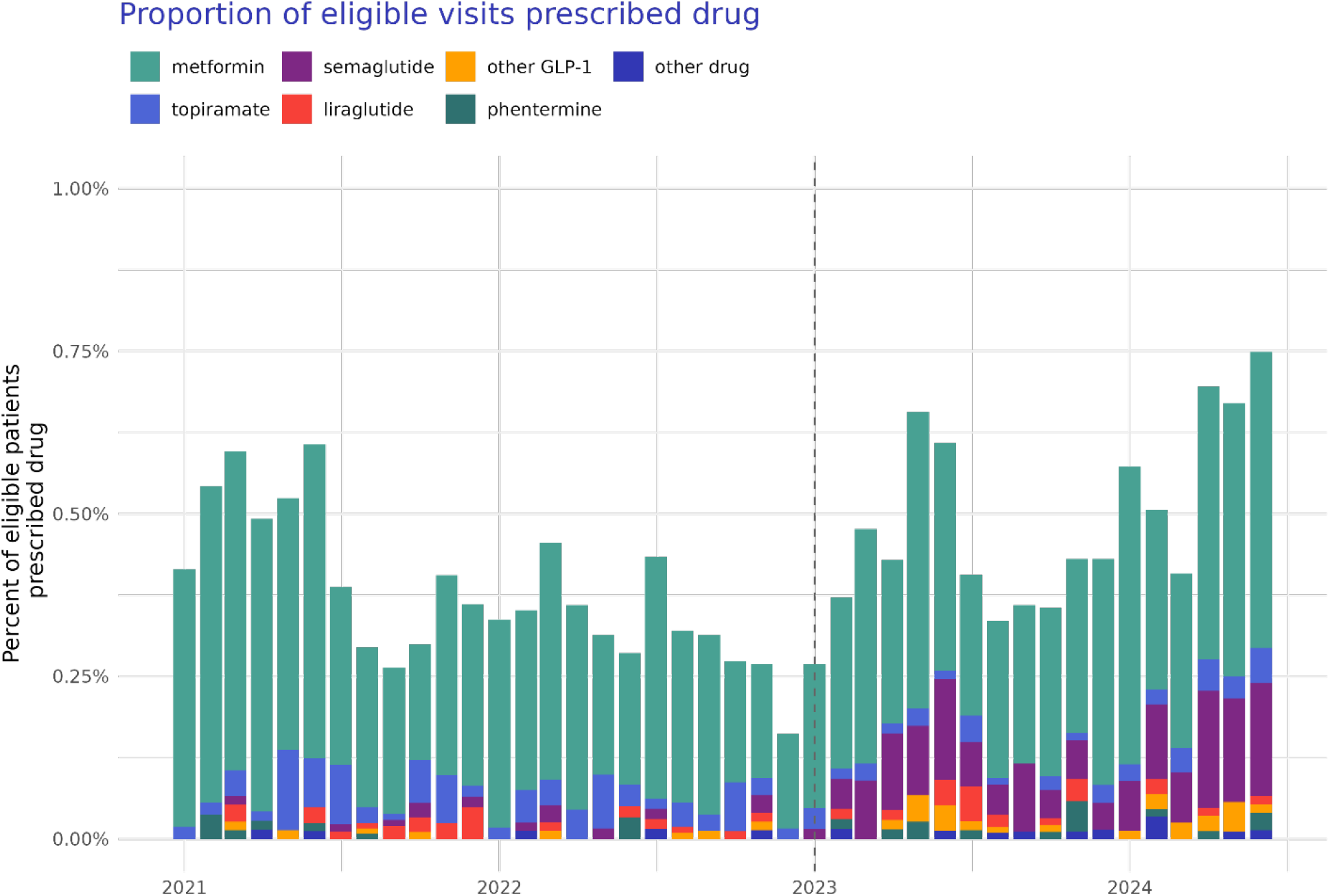
Proportion of eligible patients with that received a prescription for each medication within 14 days of their visit. Medications are not mutually exclusive. Other GLP-1s include dulaglutide, exenatide, lixisenatide, tirzepatide. Other drugs include orlistat, phentermine, and phentermine-topiramate.

## Discussion

In this large, real-world study of children and adolescents with obesity and without recent documented treatment, only a minority of patients had evidence of obesity treatment. Treatment with nutrition counseling was more common than treatment with pharmacotherapy during the entire study period. While the absolute likelihood of treatment remained low, increases in treatment levels were observed for both interventions following the AAP guideline release. Further, pharmacotherapy showed an additional increase in treatment trend since the guideline release.

These findings suggest a gradual adoption of AAP pharmacotherapy guidelines over time. However, the timing of the AAP guideline release coincided with surging national interest in GLP-1-based anti-obesity medications^20,21^ and large increases in their use across the US population.^10,14^ Therefore, changes in pharmacotherapy treatment of children and adolescents may reflect a greater acceptance of anti-obesity medications generally,^22,23^ rather than a direct impact of the AAP guidelines.

Despite early concerns about potential overuse or premature use of anti-obesity medications, we found that absolute rates of pharmacotherapy remained low in this population. Recent studies have found similarly low rates of guideline-concordant screening for obesity-related comorbidities in children and low rates of anti-obesity medication use.^13,24^ While infrequent use of pharmacotherapy is consistent across literature, estimated rates vary across studies.^13,14,25^ A recent study of youth with three or more encounters with obesity BMI between 2017 and 2020 found 8% of patients used anti-obesity medications during the period.^13^ The higher prevalence of anti-obesity medication use in that study is likely attributable to its longer assessment period, higher average BMI values, and a majority of patients taking metformin having a diagnosis of type 2 diabetes or metabolic syndrome.

Reasons for the high prevalence of nutrition counseling relative to the prevalence of pharmacotherapy in this sample may include clinician reluctance to prescribe anti-obesity medications,^26–28^ caregiver reluctance to use pharmacotherapy,^29^ and the limited insurance coverage for anti-obesity medications relative to coverage for lifestyle intervention.^30,31^ While much research is exploring policy- and systems-level opportunities to reduce the prevalence of childhood obesity,^30,32–34^ more research is needed to understand clinician-level factors including clinician-caregiver communication about obesity treatment options.^26,28,35^ This is especially relevant in the era where the social media discourse on this topic is robust and direct-to-consumer advertisement of GLP-1 medications (particularly compounded medications) is prevalent.^21,36–38^ Moreover, research about the safety profile of newer GLP-1 medications for adolescents is needed to support informed decision making about obesity pharmacotherapy.^18^

There is an extensive existing literature about barriers and facilitators to physicians offering and families engaging in childhood obesity treatment, including IHBLT and pharmacotherapy.^39,40^ Barriers include, but are not limited to, caregivers not having accurate assessments of a child’s weight class, structural barriers to behavior change and treatment engagement, lack of insurance coverage, time, insufficient clinician decision support, limited social support, and stigma. Many implementation challenges to hinder the translation of evidence-based research into practice,^41^ indicating a need for additional efforts to disseminate effective obesity treatments.

This study has several strengths. First, the data represent a large, diverse, and generalizable population of children and adolescents across the US treated in routine clinical settings. Notably, this study does not include large children’s hospitals, where rates of treatment may be higher given greater access to intensive lifestyle therapies.^31^ Unlike many studies of obesity, this study relied on BMI values measured during visits and captured in the EHR, rather than billing codes in claims data, which may be inconsistently captured.

The study is also subject to limitations. The capture of obesity treatment in EHR data is likely incomplete and may vary between health systems. Patients may have received care from other providers not represented in this dataset, and discussions about nutrition and physical activity may occur outside the formal health system or be recorded variably. Additionally, nutrition referral and counseling are proxies for IHBLT and not exclusively part of obesity treatment. However, we expect that specificity is similar in the pre- and post-guideline periods, such that misclassification is likely non-differential and would bias towards the null. Furthermore, regardless of physician intention to intervene in early obesity, a variety of barriers to effective treatment exist, including limited access to comprehensive IHBLT, geographic clustering of programs, and high costs of the most effective pharmacotherapies.^31^ High rates of unknown race in this study likely result from de-identification processes that mask certain demographic data to ensure patient privacy, rather than reflecting true unknowns among patients.^42^ Similar to other recent papers,^12^ we did not consider bariatric surgery because it typically occurs in a small subset of the adolescents at the highest BMI percentiles. Finally, we did not consider a wash-in period for the effects of the guideline change, though the inclusion of a slope term allows for gradual adoption of guidelines over time.

## Conclusion

Following new comprehensive obesity guidelines from the AAP, there were concerns about support for obesity pharmacotherapy for adolescent populations. We found that use of pharmacotherapy and nutritional counseling or referral increased significantly in children and adolescents with obesity and without T2D. Despite these large relative increases, absolute levels of obesity treatment, especially pharmacotherapy use, in this population remain low. Future research should explore clinician and family preferences for obesity treatment options, including pharmacotherapy, and understand how families balance trade-offs between treatment attributes such as time, cost, safety, uncertainty, and short- and long-term effectiveness. Implementation work is needed to improve access to these treatments. Further, consistent documentation of non-pharmaceutical treatment will aid surveillance. Continued monitoring of obesity treatment in children and adolescents is needed given rapid changes in evidence, approvals, and access to evidence-based obesity treatments.

## Data Availability

The data used in this study are available to all Truveta subscribers and may be accessed at studio.truveta.com.

