## Supplement for "Treatment of obesity in US children and adolescents before and after the AAP guidelines"

### Supplemental table 1: Codes used to identify nutrition counseling

| Code System | Concept Code | Concept Name |
|---|---|---|
| HCPCS | G0447 | Face-to-face behavioral counseling for obesity, 15 minutes |
| HCPCS | G0473 | Face-to-face behavioral counseling for obesity, group (2-10), 30 minutes |
| HCPCS | S9470 | Nutritional counseling, dietitian visit |
| ICD10CM | Z71.3 | Dietary counseling and surveillance |
| SNOMED CT | 11816003 | Diet education |
| SNOMED CT | 1230141004 | Education about nutrition influence on health |
| SNOMED CT | 1759002 | Assessment of nutritional status |
| SNOMED CT | 225388007 | Dietary intake assessment |
| SNOMED CT | 226069004 | Review of current diet |
| SNOMED CT | 226072006 | Weighed dietary intake assessment |
| SNOMED CT | 226073001 | Dietary intake assessment using food diary |
| SNOMED CT | 226074007 | Dietary intake assessment using food frequency questionnaire |
| SNOMED CT | 226075008 | Dietary intake assessment using food photographs |
| SNOMED CT | 266724001 | Weight-reducing diet education |
| SNOMED CT | 284352003 | Obesity diet education |
| SNOMED CT | 304491008 | Dietary education for disorder |
| SNOMED CT | 310243009 | Nutritional assessment |
| SNOMED CT | 370847001 | Dietary needs education |
| SNOMED CT | 410171007 | Nutrition care education |
| SNOMED CT | 410172000 | Nutrition care management |
| SNOMED CT | 410201001 | Weight maintenance regimen management |

| Code System | Concept Code | Concept Name |
|---|---|---|
| SNOMED CT | 429071001 | Dietary education for lipid disorder |
| SNOMED CT | 429072008 | Dietary education for hyperlipidemia |
| SNOMED CT | 429095004 | Dietary education for weight gain |
| SNOMED CT | 441201000124108 | Counseling about nutrition using cognitive behavioral theoretical approach |
| SNOMED CT | 441231000124100 | Counselling about nutrition using health belief model |
| SNOMED CT | 441241000124105 | Counselling about nutrition using social learning theory approach |
| SNOMED CT | 441251000124107 | Counselling about nutrition using transtheoretical model and stages of change approach |
| SNOMED CT | 441261000124109 | Counseling about nutrition using motivational interviewing technique |
| SNOMED CT | 441271000124102 | Counselling about nutrition using goal setting strategy |
| SNOMED CT | 441281000124104 | Counselling about nutrition using self-monitoring strategy |
| SNOMED CT | 441291000124101 | Counselling about nutrition using problem solving strategy |
| SNOMED CT | 441301000124100 | Counselling about nutrition using social support strategy |
| SNOMED CT | 441311000124102 | Counselling about nutrition using stress management strategy |
| SNOMED CT | 441321000124105 | Counselling about nutrition using stimulus control strategy |
| SNOMED CT | 441331000124108 | Counselling about nutrition using cognitive restructuring strategy |
| SNOMED CT | 441341000124103 | Counselling about nutrition using relapse prevention strategy |
| SNOMED CT | 441351000124101 | Counselling about nutrition using rewards and contingency management strategy |
| SNOMED CT | 443288003 | Lifestyle education regarding diet |
| SNOMED CT | 445291000124103 | Nutrition-related skill education |

| Code System | Concept Code | Concept Name |
| --- | --- | --- |
| SNOMED CT | 445301000124102 | Content-related nutrition education |
| SNOMED CT | 445331000124105 | Nutrition-related laboratory result interpretation education |
| SNOMED CT | 445641000124105 | Technical nutrition education |
| SNOMED CT | 447901000124109 | Management of nutrition-related complementary and/or alternative medicine |
| SNOMED CT | 61310001 | Nutrition education |
| SNOMED CT | 65912007 | Diet counseling |
| SNOMED CT | 699849008 | Healthy eating education |
| SNOMED CT | 710881000 | Education about eating pattern |

### Supplemental table 2: Codes used to identify nutrition referral

| Code System | Concept Code | Concept Name |
| --- | --- | --- |
| SNOMED CT | 103699006 | Refer to dietician |
| SNOMED CT | 306353006 | Referral to community-based dietitian |
| SNOMED CT | 306354000 | Referral to hospital-based dietitian |
| SNOMED CT | 408285001 | Refer to pediatric dietitian |
| SNOMED CT | 416116000 | Referral to home registered dietitian |
| SNOMED CT | 428461000124101 | Referral to nutrition professional |

### Sensitivity Analysis: Adolescents only

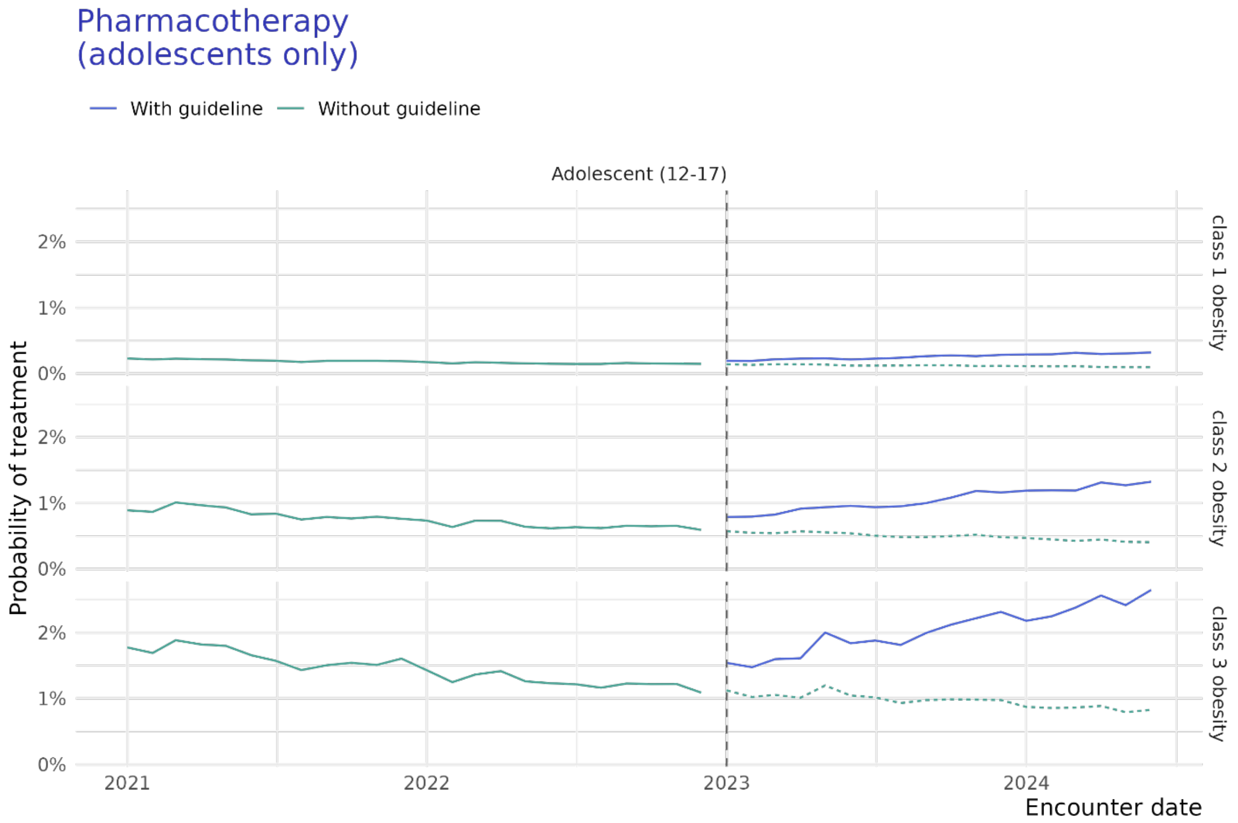


*Supplemental Figure 1: Interrupted time series of treatment with anti-obesity medication over time, from a sensitivity analysis that included adolescents only. Green lines represent the predicted probability of treatment absent guidelines, and blue lines represent predicted probabilities of treatment with the guideline, for the observed population. The dotted green line represents the counterfactual.*
